# Real world evidence of calcifediol use and mortality rate of COVID-19 hospitalized in a large cohort of 16,401 Andalusian patients

**DOI:** 10.1101/2021.04.27.21255937

**Authors:** Carlos Loucera, María Peña-Chilet, Marina Esteban-Medina, Dolores Muñoyerro-Muñiz, Román Villegas, Jose Lopez-Miranda, Jesus Rodriguez-Baño, Isaac Túnez, Roger Bouillon, Joaquin Dopazo, Jose Manuel Quesada Gomez

## Abstract

**Background:** COVID-19 is a major worldwide health problem because of acute respiratory distress syndrome, and mortality. Several lines of evidence have suggested a relationship between the vitamin D endocrine system and severity of COVID-19.

**Methods:** We present a retrospective survival study that includes all Andalusian patients hospitalized between January and November 2020 because of COVID-19 infection. Based on a central registry of electronic health records (the Andalusian Population Health Database, BPS), prescription of vitamin D or its metabolites within 15-30 days before hospitalization were recorded. The effect of treatment with vitamin D metabolites for other indication previous to the hospitalization was studied with respect to patient survival by means of Kaplan-Meyer survival curves and Log Hazard Ratios, using a propensity score to compensate the disbalance of compared classes and the confounding factors. The availability of detailed patient data in the BPS allowed to obtain Real-World Evidence (RWE) of the effects of prior use of vitamin D or its metabolites on the mortality due to COVID-19 infection.

**Findings:** A retrospective cohort of 16.401patients was extracted from the BPS, which includes all the patients hospitalized with COVID-19 diagnosis between January and November 2020 in Andalusia, one of the largest regions in Europe with the size of an average median country. A total of 358 patients were found with cholecalciferol, and 193 with calcifediol, prescriptions 15 days before hospitalization. For a period extended to 30 days before hospitalization, the numbers increase to 416 and 210 and, respectively. Kaplan-Meyer survival curves and hazard ratios support an association between consumption of these metabolites and patient survival. Such association was stronger in calcifediol (Log Hazard Ratio, LHR = -1.27±0.32) than in cholecalciferol (LHR= -0.56±0.15), when prescribed 15 days before hospitalization This effect decreases when a larger 30 days period is considered (calcifediol LHR= -1.01±0.27 and cholecalciferol LHR= -0.27±0.12), suggesting that the closer was the treatment to the hospitalization the stronger the association.

**Conclusions:** A significant reduction in mortality in patients hospitalized with COVID-19 is associated with the prescription of vitamin D, especially calcifediol, within 15-30 days prior to hospitalization.

## Introduction

Vitamin D deficiency has been associated with a large number of diseases including immune disorders and infections. The causal role of vitamin D for rickets and osteomalacia is well demonstrated and its role in aggravating osteoporosis is well accepted [1]. Vitamin D3 (cholecalciferol), the threshold nutrient of the vitamin D endocrine system (VDES), is acquired by cutaneous synthesis under the influence of UV-B light and in minimal amounts from the diet. It is transported, like other VDES metabolites, by vitamin D-binding protein (DBP) (*GC*), in the liver, mainly through the action of 25 hydroxylase (*CYP2R1* and others), where it is converted to 25OH vitamin D (25OHD) or calcifediol, biomarker of nutritional status and pre-hormone of the VDES. The 25OHD is substrate for the synthesis of 1,25(OH)2D or calcitriol through the action of 1α hydroxylase (*CYP2721B*) in the kidney, for its endocrine actions, and in multiple cells of the body for its auto/paracrine action. The system hormone calcitriol binds with high affinity to its nuclear receptor, the vitamin D receptor (*VDR*), regulating transcription of a large number (∼3%) of genes, with a broad spectrum of functional activities[1]. However, its extra-skeletal effects are more disputed. Several Mendelian Randomization studies demonstrated that genetically low serum 25-hydroxyvitamin D (25OHD) concentrations increase the risk of multiple sclerosis [2] and a meta-analysis found a reduced incidence of upper respiratory infections when supplements of vitamin D are given to relatively vitamin D deficient subjects [3]. There are now multiple association studies linking a poor vitamin D status with increased risk [4-7] or severity of COVID-19 infections [7-10] and also meta-analyses [11-13].

A recent UK study by NICE, however, concluded that there is insufficient evidence to recommend vitamin D supplementation solely for the purpose of prevention of COVID-19 (complications) but recommends the UK guidelines to prevent vitamin D deficiency in general. The NICE experts agreed that a poor vitamin D status was associated with more severe outcomes from COVID-19 but without proof of causality, especially because the risk factors for severe COVID-19 outcomes are also risk factors for low vitamin D status [14]. Similarly, a resolution of the US congress recommended correction of vitamin D deficiency in the general population as part of a global strategy to reduce the burden of COVID-19 [15]. Moreover, a recent study on systematic drug repurposing for COVID-19 based on machine learning has found that, among others, the *VDR* protein could have a protector effect over pathways affected by the SARS-CoV-2 infection [16], suggesting a potential protector role for VDES metabolites such as cholecalciferol, calcifediol or calcitriol. This study used mechanistic models [17] of the COVID-19 disease map [18] to find relevant interactions between proteins (already targets of drugs with other indications) and the pathways affected by COVID-19 disease infection either directly or downstream, collectively known as the COVID-19 disease map [18], thus providing mechanistic evidences of the protective effect of VDES metabolites in COVID-19.

Although randomized clinical trials remain the gold standard to proof efficacy and safety of whatever interventions [19], other types of studies may be faster and more efficient to provide clinical guidelines, especially when lifesaving procedures are needed in an emergency situation such as the present COVID-19 pandemic. Thus, the increasing availability of digital health data, together with the raising costs and known limitations of traditional trials, has fostered the interest in the use of real-world data (RWD) [20], defined as patient’s data on their health status and on health care received, collected from their electronic health records (EHR) [20, 21]. RWD can be analyzed to generate real word evidence (RWE) [22]. Actually, RWE provide a better image of the actual clinical environments in which medical interventions are carried out when compared to conventional randomized clinical trials, given that RWD includes detailed data on patient demographics, comorbidities, adherence, and simultaneous treatments [23, 24].

Since 2001, the Andalusian Public Health System is thoroughly storing all the EHRs data of Andalusian patients in the Population Health Base (BPS) [25]. This makes of BPS one of the largest repositories of highly detailed clinical data in the world (with over 13 million of comprehensive registries) [25]. BPS constitutes a unique and privileged environment to carry out large-scale RWE studies, with especial attention payed to the evaluation of impact in personal data protection [26].

Here we used RWD from BPS to obtain RWE of the effectiveness of the treatment of cholecalciferol, calcifediol or calcitriol VDES metabolites with nutrient, pre-hormone or hormone activity respectively previously administered by other health objectives, on mortality rate among patients hospitalized for COVID-19.

## Material and Methods

### Design and patient selection

This study aimed to study a retrospective cohort including all the Andalusian patients with COVID-19 diagnosis that were hospitalized. Following the inclusion criteria of having a COVID-19 diagnosis (by PCR or antigens test) and an overlapping hospitalization during the period January to November.

The Ethics Committee for the Coordination of Biomedical Research in Andalusia approved the study “Retrospective analysis of all COVID-19 patients in the entire Andalusian community and generation of a prognostic predictor that can be applied preventively in possible future outbreaks” (29^th^ September, 2020, Acta 09/20) and waived informed consent for the secondary use of clinical data for research purposes.

#### Data preprocessing

Medication data related to VDES metabolites in the office and hospital pharmacy records were found for the following pharmaceutical compounds: cholecalciferol, calcifediol and calcitriol. Individuals are considered as treated with a specific metabolite if prescriptions were found within a period from P days (P=15 and 30 days) before the hospital admission until the discharge (or death). Otherwise they were considered as untreated. In parallel, individuals were considered treated with All Vitamin D Metabolites (ADM) in the case that one of the previous pharmaceutical compounds were prescribed. A total of 358 patients were treated with cholecalciferol, 193 with calcifediol, and 11 with calcitriol if a period of P=15 days is considered, and 416, 210 and 15, respectively, if a P=30 days period is considered. Calcitriol was excluded from the individual studies, due to the low N, but was considered as ADM, totaling 562 and 641 for 15 and 30 days, respectively.

The main primary outcome used here was COVID-19 death events (certified death events during hospitalization). Following previous similar studies, the first 30 days of hospital stay were considered for survival calculations [27]. The time variable in the models correspond to the length (in days) of hospital stay.

#### Propensity score matching

To reduce the confounding effects of several conditions on the outcome a 1:1 ratio Propensity Score Matching (PSM) was applied to match treated and untreated patients without replacement in the survival analysis. Variables previously associated with COVID-19 mortality, such as: age, sex, pneumonia/flu vaccination status, hypertension, chronic obstructive pulmonary disease, diabetes, obesity, chronic pulmonary and digestive diseases, asthma, chronic heart diseases and cancer were included [28] (Table 2). The propensity scores have been estimated by means of a Generalized Additive Model with a logit as the link function while the matching, to ensure a similar distribution of all the covariates across treatment groups, has been done using the nearest neighbor matching modality [29]. To ensure a similar distribution of all the covariates across treatment groups (beyond considering only standardized means differences) the higher order moments of covariates were used as recently recommended [30]. Covariate balance in matched samples is checked by a X^2^ test to confirm that no biases against treated or untreated matched patients exist.

#### Survival on the matched samples

Kaplan-Meier estimate was used to infer the survival probability difference between treated and untreated patients. Survival curves for the different treatments are compared with a Log Rank test.

#### Robust estimation of the treatment effect using the whole population

Although PSM is a widely used technique because it leverages the use of parametric and non-parametric models to covariate-treatment-outcome unbalanced data, the consistency of any estimator derived from the propensity scores is limited by exchangeability assumptions between the treated and untreated samples, the covariate adjustment and model specification (among others) mainly due to the fact that the propensity score is computed with the same data as the modelling. Here, the hazard ratios for each of the treatments of interest have been computed by means of the closed-form estimator [31] using a weighted Cox model with inverse propensity weighting under the Average Treatment Effect on the Overall (ATE) and the Average Treatment Effect on the Treated (ATT) assumptions, the most used weighting approximations to estimate treatment effects [32]. Note that the ATE weights are stabilized by factoring the overall probability of being exposed to a given treatment into the equation [31].

Furthermore, an alternative estimation of the treatment effect has been obtained by means of bootstrapping (n=10000 iterations) a weighted Cox model with the propensity weights computed by means of a Binomial General Linear Model (GLM) which regress the treatment as a function of the covariates [33].

#### Modeling survival along time

For each time point the restricted mean survival time (RMST) of the treated versus the untreated for each treatment has been compared. The RMST is computed as the area under the survival curve up to the time point (*t*) and, therefore, the comparison measures the difference and ratio of RMST between treated and untreated patients. The interpretation of the curve is straightforward, representing each time point (*t*) the expected days (on average) that subjects from the treatment group live longer (or shorter) than untreated patients when patients are followed up to time *t*. Interestingly, the significance of the RMST comparison can be estimated (FDR corrected p-value) for each time point. Note that the dynamic estimated ratio of RMST is more prone to detect plateaus on the treatment effect over time [34], so both curves are complementary.

#### Software

For the matching analysis we have used the *MatchIt* [35] R package (version 4.1.0). The treatment effect models have been implemented with the *hrIPW* [36] R package (version 0.1.3). RMST computations have been performed with the survRM2 [37] R package (version 1.0.3). Survival curves and plots have been generated with the R *survival* [38] (version 3.2.7) and *survminer* [39] (version 0.4.8) packages, respectively.

## Results

### Data processing

A retrospective cohort of 16.401 patients, which include all Andalusian patients with COVID-19 diagnosis that were hospitalized between January and November 2020, was found in BPS and collected. Supplementary Figure 1 depicts the frequency of hospital admission of patients with COVID-19 diagnosis along this period. Patient data on medication and other relevant covariates (see Table 1) was downloaded from the BPS.

**Table 1.** Data imported from BPS for each patient

| **Code** | **Meaning** |
| --- | --- |
| FECNAC | Birth date |
| FECDEF | Death date |
| SEXO | Gender |
| FEC_INGRESO | Hospital admission date |
| FEC_ALTA | Discharge date |
| MOTIVO_ALTA | Reason for the discharge: (recovery/death/admission in another hospital/voluntary discharge/retirement home/unspecified) |
| DIAS_UCI | Days in ICU |
| COD_PATOLOGIA_CRONICA | Hospital codes for chronic conditions |
| COD_FEC_INI_PATOLOGIA | Date of condition diagnosis |
| COD_CIE_NORMALIZADO | A mixture of ICD9 and ICD10 codes for diseases |
| DESC_CIE_NORMALIZADO | Description of the ICD |
| FECINI_DIAG | Diagnosis date |
| FECFIN_DIAG | End of the diagnosed condition |
| FUENTE_DIAG | Source of the diagnosis (hospital, emergency, etc.) |
| IND_CRONICO_HCUP | Is a chronic disease? (yes/no) |
| Test COVID: FECHA | Test COVID date |
| Test COVID: TYPE | PCR / antigens |
| Test COVID: RESULTADO_TEST | Result of the test (positive/negative)<sup>1</sup> |
| Pharmacy (Hospital and external): DESCRIPCION | List of drugs used in hospital or purchased in the pharmacies |
| Pharmacy (Hospital and external): FECHA | Dispensing date |
| VACUNA | List of vaccines |
| VACUNAFECHA | Vaccination dates |

<sup>1</sup> Only positive were considered in this study

**Table 2.** Variables previously associated to COVID-19 prognosis and symptoms observed in the whole set of patients. Columns survival and death contain the absolute number of individuals with the specific covariate, and in parentheses the percentage, that survive and die, respectively. A X^2^ test is carried out to check for direct associations (with no covariate correction) with death. The difference of percentages accounts for the effect: e.g. asthma protects and cancer increases the risk. The p-value column accounts for the significance. All the values agree with previous reports.

| Days | Covariate | Survival | Death | p-value |
| --- | --- | --- | --- | --- |
| 30 | Total N | 13350 | 3019 |  |
| 30 | N cross-treated | 27 | 1 |  |
| 30 | Asthma | 1642 (12.3) | 291 (9.6) | <0.001 |
| 30 | Cancer | 1513 (11.3) | 630 (20.9) | <0.001 |
| 30 | Chronic heart diseases | 3318 (24.9) | 1397 (46.3) | <0.001 |
| 30 | Diabetes | 3788 (28.4) | 1319 (43.7) | <0.001 |
| 30 | Hypertension | 7537 (56.5) | 2306 (76.4) | <0.001 |
| 30 | Obesity | 2320 (17.4) | 517 (17.1) | 0.760 |
| 30 | Chronic pulmonary diseases | 1750 (13.1) | 778 (25.8) | <0.001 |
| 30 | Flu vaccine | 5654 (42.4) | 2003 (66.3) | <0.001 |
| 30 | Pneumococcal vaccine | 3481 (26.1) | 1241 (41.1) | <0.001 |
| 30 | Dementia | 990 (7.4) | 588 (19.5) | <0.001 |
| 30 | Sex (Female) | 6203 (46.5) | 1241 (41.1) | <0.001 |
| 30 | Age_bin |  |  | <0.001 |
| 30 | (-0.001, 41.0] | 1671 (12.5) | 34 (1.1) |  |
| 30 | (41.0, 68.0] | 6094 (45.6) | 505 (16.7) |  |
| 30 | (68.0, 105.0] | 5585 (41.8) | 2480 (82.1) |  |
| 15 | Total N | 13354 | 3020 |  |
| 15 | N cross treated | 23 | 0 |  |
| 15 | Asthma | 1645 (12.3) | 291 (9.6) | <0.001 |
| 15 | Cancer | 1513 (11.3) | 630 (20.9) | <0.001 |
| 15 | Chronic heart diseases | 3319 (24.9) | 1398 (46.3) | <0.001 |
| 15 | Diabetes | 3790 (28.4) | 1320 (43.7) | <0.001 |
| 15 | Hypertension | 7541 (56.5) | 2307 (76.4) | <0.001 |
| 15 | Obesity | 2320 (17.4) | 517 (17.1) | 0.759 |
| 15 | Chronic pulmonary diseases | 1752 (13.1) | 778 (25.8) | <0.001 |
| 15 | Flu vaccine | 5658 (42.4) | 2004 (66.4) | <0.001 |
| 15 | Pneumococcal vaccine | 3482 (26.1) | 1241 (41.1) | <0.001 |
| 15 | Dementia | 990 (7.4) | 589 (19.5) | <0.001 |
| 15 | Sex (Female) | 6207 (46.5) | 1242 (41.1) | <0.001 |
| 15 | Age_bin |  |  | <0.001 |
| 15 | (-0.001, 41.0] | 1671 (12.5) | 34 (1.1) |  |
| 15 | (41.0, 68.0] | 6096 (45.6) | 505 (16.7) |  |
| 15 | (68.0, 105.0] | 5587 (41.8) | 2481 (82.2) |  |

### Vitamin D endocrine system metabolites and survival

The effect of cholecalciferol, calcifediol or calcitriol treatment, both aggregated (ADM) and independently, 15 and 30 days before hospitalization, was studied with respect to the outcome death at 30 days. As described in methods, PSM was applied to the treated and untreated patients. This rendered a satisfactory covariate balance and no significant correlations between the covariates was observed in the samples paired by the PSM model (Table 3). Kaplan-Meier curves shows the survival of patients treated with ADM 15 days (Figure 1A) and 30 days (Figure 1B) before hospitalization, suggesting a significant association between ADM treatment and patient survival. Kaplan-Meier curves for specific cholecalciferol, calcifediol or calcitriol treatments (Supplementary Figure 2) supporting the same significant association between any of the individual treatments and patient survival. The comparison of specific treatments supports a significantly increased survival of patients treated with calcifediol than those treated with cholecalciferol (see Table 4), pointing to a stronger association of calcifediol with patient survival.

**Table 3.** Matched covariates across treated and untreated patients. A X^2^ test (column p-value) systematically non-significant demonstrates that the values are equilibrated between both groups.

| Days | Covariate | ADM Treated | ADM Untreated | p-value |
| --- | --- | --- | --- | --- |
| 30 | Total N | 641 | 641 |  |
| 30 | Asthma | 83 (12.9) | 63 (9.8) | 0.095 |
| 30 | Cancer | 105 (16.4) | 88 (13.7) | 0.211 |
| 30 | Chronic heart diseases | 220 (34.3) | 226 (35.3) | 0.769 |
| 30 | Diabetes | 200 (31.2) | 198 (30.9) | 0.952 |
| 30 | Hypertension | 412 (64.3) | 417 (65.1) | 0.815 |
| 30 | Obesity | 104 (16.2) | 104 (16.2) | 1.000 |
| 30 | Chronic pulmonary diseases | 108 (16.8) | 93 (14.5) | 0.282 |
| 30 | Flu vaccine | 353 (55.1) | 356 (55.5) | 0.911 |
| 30 | Pneumococcal vaccine | 241 (37.6) | 249 (38.8) | 0.687 |
| 30 | Dementia | 50 (7.8) | 47 (7.3) | 0.833 |
| 30 | Sex (Female) | 398 (62.1) | 397 (61.9) | 1.000 |
| 30 | Age_bin |  |  | 0.321 |
| 30 | (-0.001, 41.0] | 41 (6.4) | 54 (8.4) |  |
| 30 | (41.0, 68.0] | 239 (37.3) | 224 (34.9) |  |
| 30 | (68.0, 105.0] | 361 (56.3) | 363 (56.6) |  |
| 15 | Total N | 562 | 562 |  |
| 15 | Asthma | 76 (13.5) | 65 (11.6) | 0.368 |
| 15 | Cancer | 90 (16.0) | 75 (13.3) | 0.238 |
| 15 | Chronic heart diseases | 181 (32.2) | 181 (32.2) | 1.000 |
| 15 | Diabetes | 171 (30.4) | 165 (29.4) | 0.745 |
| 15 | Hypertension | 359 (63.9) | 366 (65.1) | 0.708 |
| 15 | Obesity | 85 (15.1) | 75 (13.3) | 0.442 |
| 15 | Chronic pulmonary diseases | 92 (16.4) | 75 (13.3) | 0.180 |
| 15 | Flu vaccine | 299 (53.2) | 306 (54.4) | 0.720 |
| 15 | Pneumococcal vaccine | 191 (34.0) | 194 (34.5) | 0.900 |
| 15 | Dementia | 43 (7.7) | 38 (6.8) | 0.645 |
| 15 | Sex (Female) | 350 (62.3) | 347 (61.7) | 0.902 |
| 15 | Age_bin |  |  | 0.764 |
| 15 | (-0.001, 41.0] | 37 (6.6) | 42 (7.5) |  |
| 15 | (41.0, 68.0] | 220 (39.1) | 225 (40.0) |  |
| 15 | (68.0, 105.0] | 305 (54.3) | 295 (52.5) |  |

**Table 4.** Comparison between the survival curves by the Log Rank test with the corresponding p-values both unadjusted and FDR-adjusted.

| Days | Treatment | Calcifediol<br>(FDR-adjusted) | Cholecalciferol<br>(FDR-adjusted) | Calcitriol<br>(FDR-adjusted) | Calcifediol<br>(unadjusted) | Cholecalciferol<br>(unadjusted) | Calcitriol<br>(unadjusted) |
| --- | --- | --- | --- | --- | --- | --- | --- |
| 30 | Cholecalciferol | 0,0270* | - | - | 0,0090** | - | - |
| 30 | Calcitriol | 0,7253 | 0,8488 | - | 0,4835 | 0,8488 | - |
| 30 | Untreated | 0,0030** | 0,4320 | 0,8488 | 0,0005** | 0,2160 | 0,8211 |
| 15 | Cholecalciferol | 0,1299 | - | - | 0,0649 | - | - |
| 15 | Calcitriol | 0,8080 | 0,8268 | - | 0,6733 | 0,8268 | - |
| 15 | Untreated | 0,0074** | 0,1299 | 0,6511 | 0,0012** | 0,0562 | 0,4341 |

\* Test significant at $\alpha < 0.05$ ; \*\* test significant at $\alpha < 0.01$ ;

**Figure 1.**
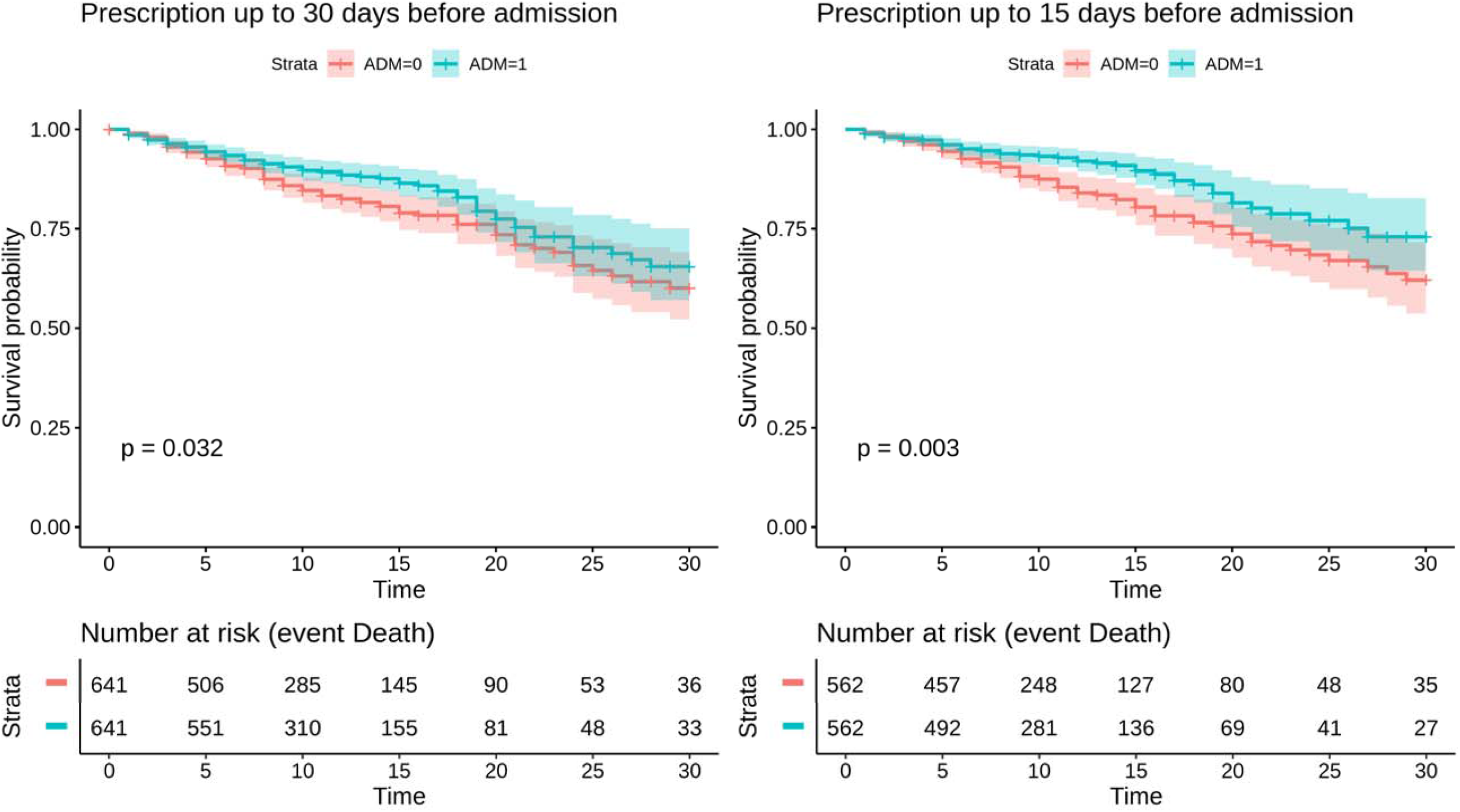
Kaplan-Meier curves of patients treated with ADM A) 15 days and B) 30 days before hospitalization for death outcome.

To study in more detail the protective effect of VDES treatment a Cox regression was used to estimate the degree of association between the treatments and death risk by means of the hazard ratios. Figure 2 summarizes the log hazard ratios with respect to the outcome death for the calcifediol and cholecalciferol treatments as well as the ADM treatments aggregated, in the two periods of administration considered (15 and 30 days). The three treatments demonstrated a significant association with increased patient survival. From Figure 2 it becomes apparent that calcifediol shows a clearly higher association with patient survival than cholecalciferol, although this last metabolite also shows a non-negligible and significant effect. Calcitriol was not included, given the small sample size (16 patients). Additional analysis with different assumptions and different methodologies, such as bootstrap) support the results obtained (Supplementary Figure 3).

**Figure 2.**
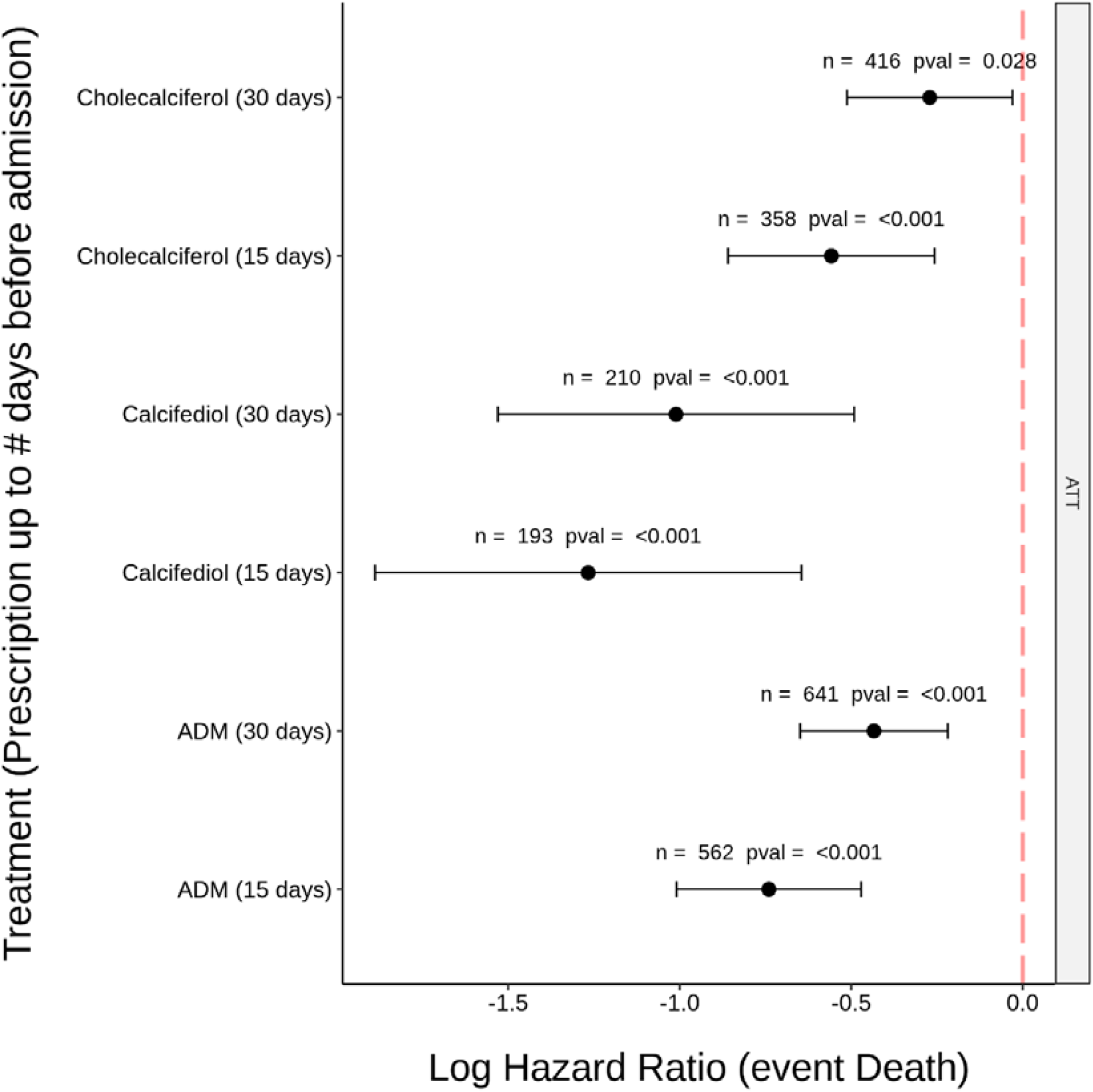
Log hazard ratios with respect to the outcome death under ATT assumption for the cholecalciferol, calcifediol and ADM treatments in the two periods considered (15 and 30 days before hospitalization). In all the cases a significant protective effect is observed: confidence intervals do not cross the 0 line (_α_=0.05)

Additionally, a more sophisticated analysis has been carried out to determine in detail the effect of each treatment along time (RMSTs). The RMST curve represents the expected survival days (on average) that subjects from the treatment group have with respect to untreated patients along time. Supplementary Figure 4 shows that calcifediol treatment shows a significantly better survival than untreated for all the 30 days interval studied. Cholecalciferol treatment shows better survival than untreated patients as well, although it is only statistically significant for a short period, and only in the case of prescription during the 15 days before admission.

## Discussion

Andalusia, with 8.5 million inhabitants is the third largest region in Europe, having a population similar to Austria and being bigger than half of the countries of the European Union. Moreover, it has the largest population under a universal EHR. All the data recorded by the Andalusian Public Health System is stored in the BPS. This allowed an unprecedented region-wise cohort study of 16,401 patients, which corresponds to all the COVID-19 patients that were hospitalized between January and November 2020.

This large-scale RWE study clearly shows that pre-treatment for another indication with VDES metabolites significantly reduces the risk of death in patients hospitalized for COVD-19, This effect is stronger in patients taking calcifediol, but also occurs in patients taking cholecalciferol and most likely with calcitriol as well (although due to the small sample size its effect was not statistically significant). To our knowledge, this is the first study to investigate the effect of pre-admission treatment with VDES metabolites (cholecalciferol, calcifediol or calcitriol) on death in patients hospitalized for COVID. Moreover, due to the country-size scale of this observational study it is easy to mimic the randomization element of an RTC and properly compare treatment groups, given the number of individuals available to properly adjust for all baseline cofounders [40]. Actually, the use of propensity scores provides additional adjustment to control for confounding variables [41]. Here, as shown in Table 3, confounding effects between the compared groups due to the known variables associated to the outcomes considered can be ruled out.

The consistency of the results presented here strongly suggests that patients that had adjusted their serum levels of 25OHD for other health objectives presented a better response to COVID-19. Recently, numerous epidemiological and association studies have been published investigating the links between circulating levels of 25OHD, and the incidence and severity of COVID-19 infections [6-13]. Initially, these were observational ecologic studies that described a higher incidence of COVID-19 infection and death in countries where vitamin D deficiency (or low sun exposure) was common [11, 42-44]. Thereafter, several studies evaluated the association between vitamin D status and risk or severity of SARS-CoV-2 infection [4-11]. These results suggest that improving serum 25OHD concentration may improve the prognosis of COVID-19 [11-13], as demonstrated by a pilot controlled trial that reported that administration of calcifediol versus no calcifediol reduced the need for ICU treatment in 76 hospitalized participants with COVID-19 who also received best available therapy [45, 46].

From a mechanistic perspective, there is good reason to postulate that the vitamin D endocrine system may have beneficial effects on different stages of COVID-19 infections such as the early viral infection (by innate antiviral effector mechanisms, including induction of antimicrobial peptides and autophagy) and the later hyperinflammatory phase of COVID-19 [47-49]. Moreover, activation of the vitamin D receptor (*VDR*) signaling pathway may have a critical modulatory role to host responses in acute respiratory distress syndrome [1, 48] by decreasing the cytokine/chemokine storm, producing a shift towards amplified adaptive Th2 immune responses, regulating the renin-angiotensin-bradykinin system (RAS), modulating neutrophil activity [50] and maintaining the integrity of the pulmonary epithelial barrier, stimulating epithelial repair [51-53] and decreasing the increased coagulability and prothrombotic tendency associated with severe COVID-19 [47, 48, 54, 55] Regulation of the renin-angiotensin-bradykinin system is of particular relevance in mitigating the progression of severe COVID-19, where over-activation of RAS is associated with a poor prognosis [56]. Moreover, the protective effect of drugs targeting the *VDR* and the *GC* (Vitamin D Binding Protein or *VDBP*) proteins of VDES has been suggested in a recent study on systematic drug repurposing for COVID-19 [16]. The ML study has demonstrated the relevance of drugs targeting *VDR* and *GC* (*VDBP*) proteins in the activity of COVID19-related signaling circuits (see Supplementary Table 1). These signaling circuits affect cellular processes involved in modulating the immune activity, decreasing the inflammatory response, but also in slowing down cellular energetics.

Thus, both observational evidence and mechanistic knowledge support a crucial role of the vitamin D endocrine system, especially calcifediol, in the response to severe outcomes of the COVID-19.

## Conclusions

This study strongly suggests that calcifediol or cholecalciferol treatments established previously to hospitalization were associated with a better survival rate among patients hospitalized because of COVID-19, most likely through *VDR* stimulation. VDES metabolite treatment may represent an effective, accessible, safe, well-tolerated and cost-effective preventive therapeutic approach for COVID-19, which is dramatically increasing in incidence and for which few validated treatments currently exist. Further large prospective, preferably interventional, Randomized Controlled Trials are needed to confirm whether regular treatment or supplementation of older adults with calcifediol or vitamin D3 improves COVID-19 outcomes.

The results reported here support the establishment of public health policies that make it possible to maintain adequate levels of 25OHD for the synthesis of calcitriol to enable a better prognosis in patients affected by COVID-19. In the light of the results obtained, calcifediol preferably, or cholecalciferol with a lower effect, can adequately meet these objectives. In fact, calcifediol may have some advantages over native vitamin D3. Thus, the former has a more reliable intestinal absorption (close to 100 %) and can rapidly restore serum concentrations of 25OHD as it does not require hepatic 25-hydroxylation. This is especially relevant in clinical situations whereby rapid restoration of serum 25OHD is desirable and *CYP2R1* expression is compromised [57]. This cost-effective and widely available treatment could have positive implications for the management of COVID-19 worldwide, particularly in developing countries.

## Data Availability

No data were generated in this study

## Authors’ contributions

Carlos Loucera: Data curation, Formal Analysis, Investigation, Visualization; María Peña-Chilet: Formal Analysis; Marina Esteban-Medina: Formal Analysis; Dolores Muñoyerro-Muñiz: Resources; Román Villegas: Resources; Jose Lopez-Miranda: Writing – review & editing; Jesus Rodriguez-Baño: Writing – review & editing; Isaac Túnez: Writing – review & editing; Roger Bouillon: Writing – review & editing; Joaquin Dopazo: Supervision, Project administration, Writing – original draft; Jose Manuel Quesada Gomez: Supervision, Writing – original draft.

## Ethics committee approval

The Ethics Committee for the Coordination of Biomedical Research in Andalusia approved the study “Retrospective analysis of all COVID-19 patients in the entire Andalusian community and generation of a prognostic predictor that can be applied preventively in possible future outbreaks” (29th September, 2020, Acta 09/20) and waived informed consent for the secondary use of clinical data for research purposes.

## Acknowledgements

This work is supported by grants SAF2017-88908-R from the Spanish Ministry of Economy and Competitiveness, PT17/0009/0006, ACCI2018/29 from CIBERER-ISCIII; COV20/00788 from the Instituto de Salud Carlos III (ISCIII), co-funded with European Regional Development Funds (ERDF); grant G999088Q from the Fundación BBVA; grant H2020 Programme of the European Union grants Marie Curie Innovative Training Network “Machine Learning Frontiers in Precision Medicine” (MLFPM) (GA 813533); P18-RT-3471 from Consejería de Salud y Familias de la Junta de Andalucía; CB16/10/00245, CB16/10/00501 from CIBERFES-ISCIII; PI19/00033 from the ISCIII, co-funded with ERDF; COVID-011-2020 from Consejería de Salud y Familia. The authors also acknowledge Junta de Andalucía for the postdoctoral contract of Carlos Loucera (PAIDI2020-DOC_00350) co-funded by the European Social Fund (FSE) 2014-2020.

