## Supplementary Figure 1 for "Real world evidence of calcifediol use and mortality rate of COVID-19 hospitalized in a large cohort of 16,401 Andalusian patients"

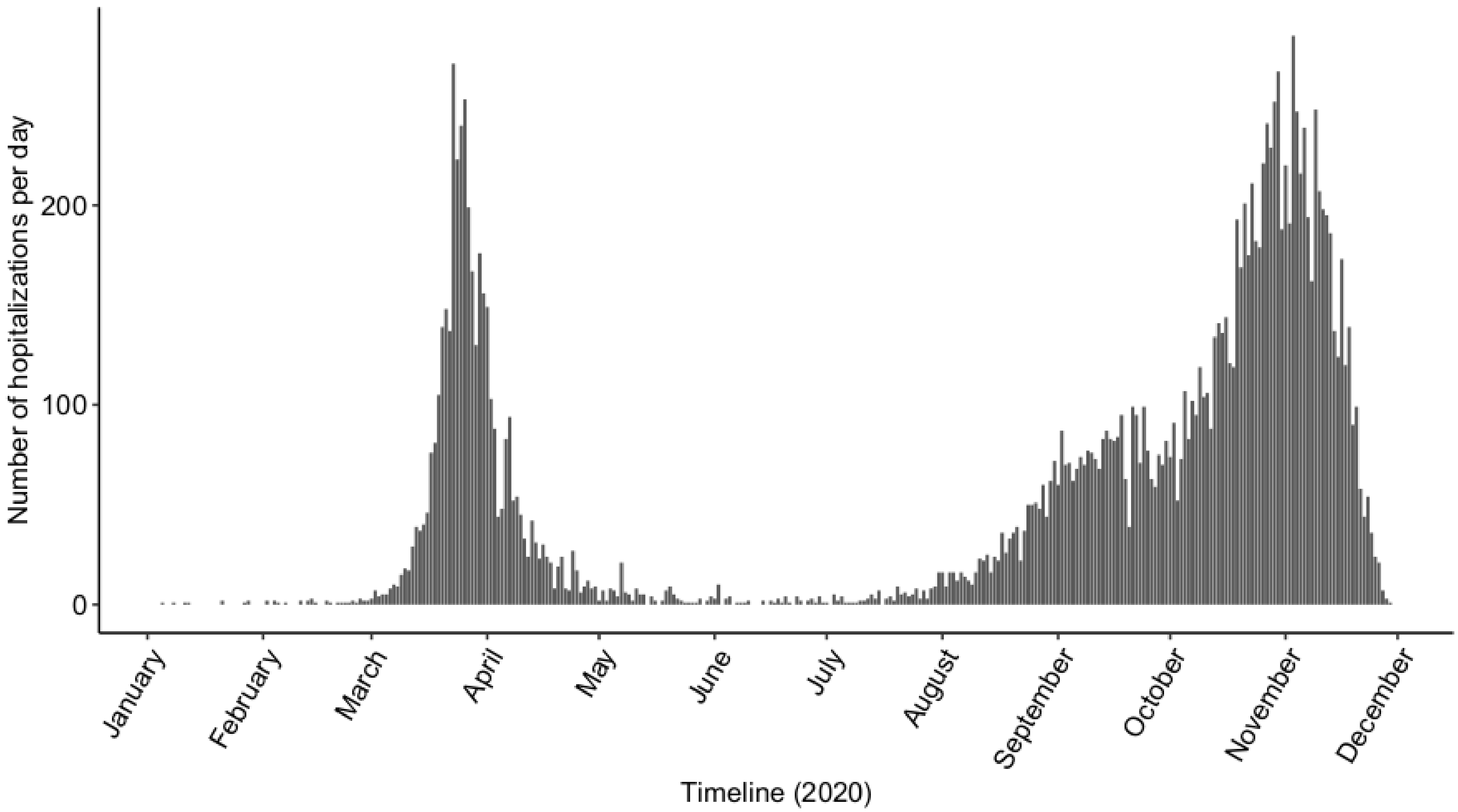


**Supplementary Figure 1**. Hospitalization dates of the patients included in the study along the period studies (January to November, 2020). The admission trend is clearly overlapping with the COVID-19 waves. Obviously, for a few early hospitalizations the diagnosis was posterior.
