## Supplementary Figure 2 for "Real world evidence of calcifediol use and mortality rate of COVID-19 hospitalized in a large cohort of 16,401 Andalusian patients"

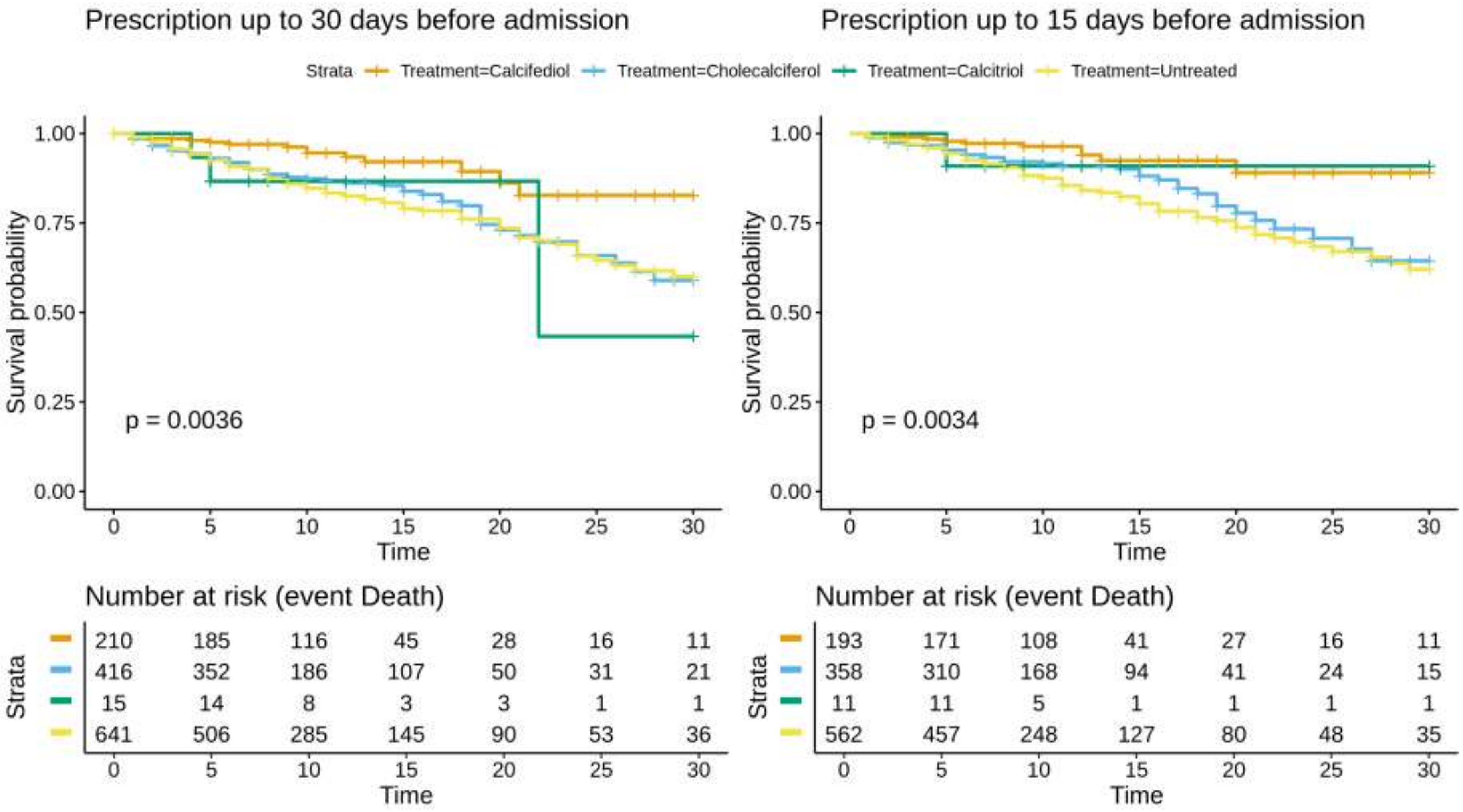


**Supplementary Figure 2. Kaplan-Meier curve for calcifediol, cholecalciferol and calcitriol for death outcome.**
