## Supplementary Figure 3 for "Real world evidence of calcifediol use and mortality rate of COVID-19 hospitalized in a large cohort of 16,401 Andalusian patients"

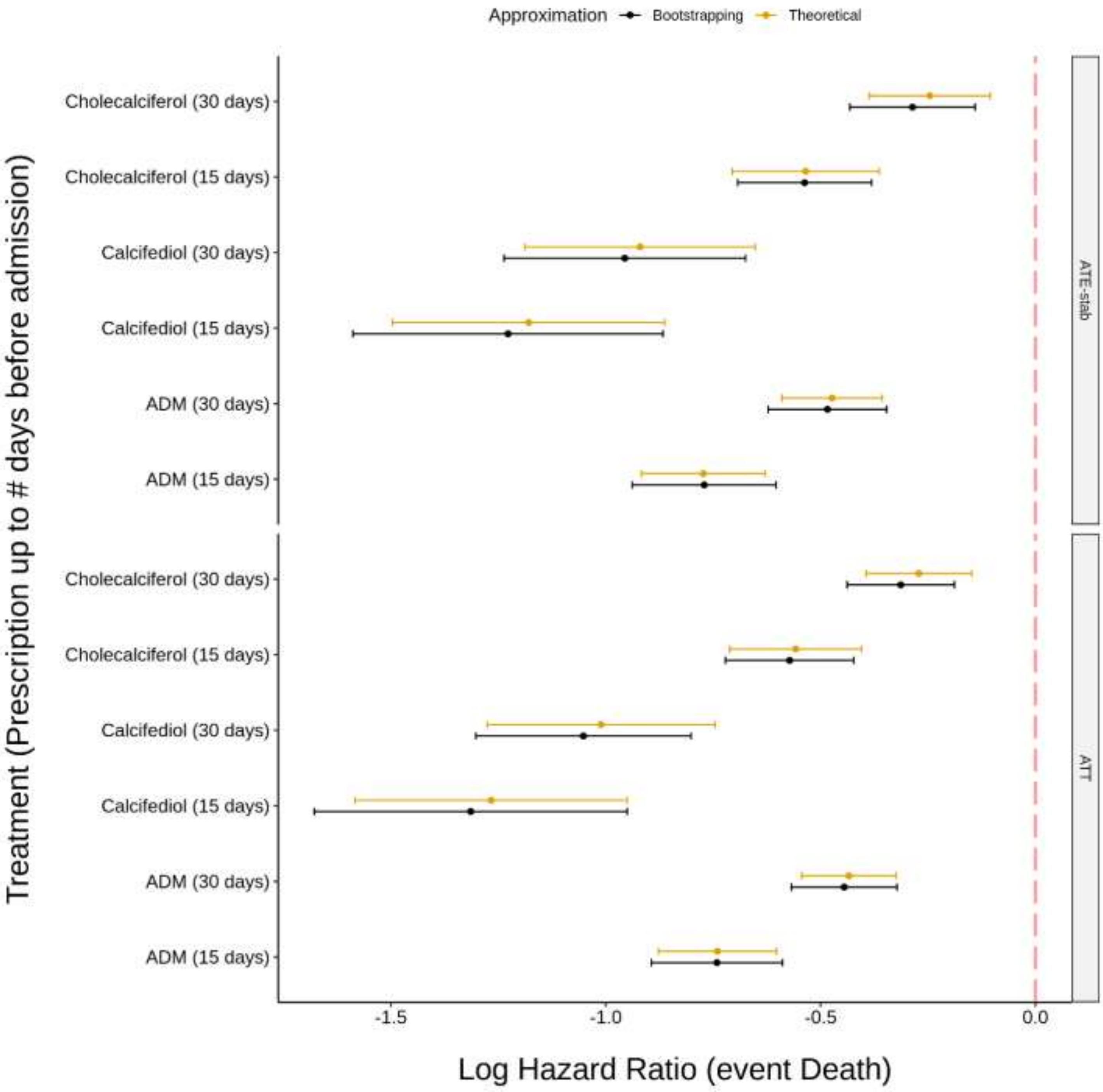


**Supplementary Figure 3**. Bootstrapping (n=10000 iterations) of a Cox model with the propensity weights computed by means of a Binomial General Linear Model (GLM) which regress the treatment as a function of the covariates. The bootstrapping approximates the hazard estimations of the closed form under both estimands: ATE and ATT for outcome: death.
