## Supplementary Figure 4 for "Real world evidence of calcifediol use and mortality rate of COVID-19 hospitalized in a large cohort of 16,401 Andalusian patients"

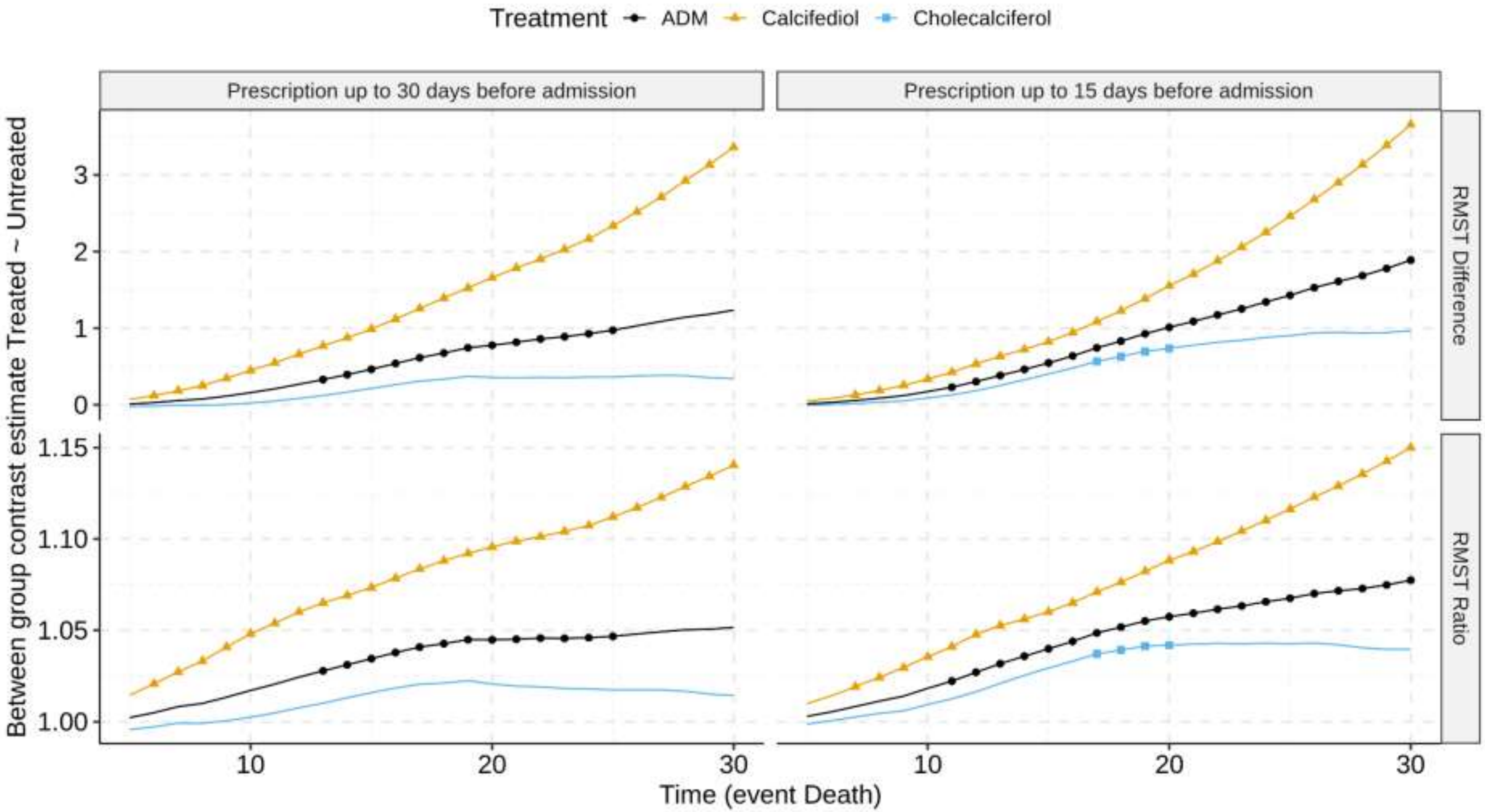


**Supplementary Figure 4. RMST curves for the three treatments**. RMST represents the expected survival days (on average) that subjects from the treatment group have with respect to untreated patients along time both as number of days (upper panels) or as a ratio (lower panels) for both prescriptions 15 days before admission (right panels) and 30 days (left panels). Dots (triangles, squares or circles) in the curves correspond to time points with a significant RMST, FDR-adjusted p-value < 0.05.
