## Supplementary Table 1 for "Real world evidence of calcifediol use and mortality rate of COVID-19 hospitalized in a large cohort of 16,401 Andalusian patients"

**Supplementary Table 1**. Signaling circuits found to be relevantly affected (according to the ML algorithm) by the *VDR* and *GC* genes, which are targets of calcifediol, cholecalciferol and calcitriol. Signaling circuits are sub-pathways defined within KEGG pathways. They are named as “pathway:gene”, being the pathway to which they belong to and the effector gene at the end of the signal transduction circuit that triggers the function(s) defined by the GO terms and summarized as general COVID-19 hallmarks. These signaling circuits affect cellular processes involved in modulating the immune activity and decreasing inflammatory response, and also in slowing down the cellular energetics. All these processes have been described in COVID-19 and linked with a higher susceptibility to develop severe symptoms.

| **Circuit** | ***VDR* relevant** | ***VDR* score** | ***GC* relevant** | ***GC* score** | **COVID-19 Hallmarks** | **GO function (from UniProt)** |
|---|---|---|---|---|---|---|
| Complement and coagulation cascades: *C2* | - | - | Y | 6,88E-04 | Immune activity | Complement pathway, Innate immunity, Immunity |
| Inflammatory mediator regulation of TRP channels: *TRPM8* | - | - | Y | 7,06E-04 | Energetics, Inflammatory response | Ion channel, Ion transport, Transport, Sensory transduction |
| Insulin signaling pathway: *PKLR* | - | - | Y | 2,50E-04 | Energetics | Glycolysis |
| Insulin signaling pathway: *GYS1* | - | - | Y | 1,04E-04 | Energetics | Gluconeogenesis |
| Adipocytokine signaling pathway: *PTPN11* | Y | 5,98E-04 | - | - | Inflammatory response | abortive mitotic cell cycle [GO:0033277]; activation of MAPK activity [GO:0000187]; cellular response to cytokine stimulus [GO:0071345]; epidermal growth factor receptor signaling pathway [GO:0007173]; fibroblast growth factor receptor signaling pathway [GO:0008543]; T cell co-stimulation [GO:0031295] |
| Adipocytokine signaling pathway: *POMC* | Y | 5,51E-04 | - | - | Immune activity, Anti-viral defense | antimicrobial humoral immune response mediated by antimicrobial peptide [GO:0061844]; cell-cell signaling [GO:0007267]; killing of cells of other organism [GO:0031640] |
